# Resting State Functional MRI Connectivity Association with Consciousness, Mortality, Longitudinal and Two-Year Outcomes in Neonatal Acute Brain Injury

**DOI:** 10.1101/2022.06.07.22275838

**Authors:** Varina L. Boerwinkle, Bethany L. Sussman, Iliana Manjón, Alyssa McGary, Mirea Lucia, Jordan Broman-Fulks, Senyene Hunter, Sarah Wycoff, Kim Allred, Deborah Tom

## Abstract

**Background:** An accurate and comprehensive test of integrated brain network function is needed for neonates during the acute brain injury period to inform on morbidity. In our first term neonatal acute brain injury (ABI) study we demonstrated resting state functional MRI (RS) acquired within 31 days of life, results in disrupted connectivity of the resting state fMRI networks, incrementally associated with consciousness, mortality, cognitive and motor development, and ongoing concern for seizures at 6 months post-gestation. In this retrospective cohort study, we evaluate extended 2-year outcomes in the same patients.

**Methods:** Study subjects included the same 40 consecutive neonates from our prior study, with resting state functional MRI acquired within 31 days after suspected brain insult from March 2018 to July 2019. Acute-period exam and test results were assigned ordinal scores based on severity as documented by respective treating specialists. Analyses (Fisher exact, Wilcox Sum-Rank test ordinal/multinomial logistic regression) examined association of resting state networks with demographics, presentation, neurological exam, electroencephalogram, anatomical MRI, magnetic resonance spectroscopy, passive task functional MRI, and outcomes of NICU and all mortality, outpatient development measured by exam and the Pediatric Cerebral Performance Category Scale (PCPC), motor development and tone, and ongoing concern for seizure at up to 42 months of age. All statistical tests were 2-sided, with statistical significance and CI adjusted using a Bonferroni correction to account for multiple test comparisons for each network and other modality.

**Results:** Subjects had a mean (standard deviation) gestational age of 37.8 (2.6) weeks, follow-up median age follow-up median age (interquartile range) 30.5 (23.6, 36.7) months, 68% were male, with a diagnosis of hypoxic ischemic encephalopathy (60%). Of the 40 patients, three died prior to discharge, and another four between 6-42 months, and 5 were lost to follow-up. Of the followed, findings at birth included mild distress (46%), moderately abnormal neurological exam (34%), and consciousness characterized as awake but irritable (37%). Significant associations after multiple testing corrections were detected for resting state networks: basal ganglia with PCPC (odds ratio [OR], 9.54; 99.4% confidence interval [CI], 1.89-48.1; P = 0.0003), NICU mortality (OR, 57.5; 99% CI, 1.35->999; P = 0.006), outpatient mortality (OR, 65.7; 99% CI 1.47->999; P = 0.005), and motor tone/weakness (OR, 17.8; 99% CI, 2.2-143; P = 0.0004); language/frontoparietal network with developmental delay (OR, 3.64; 99% CI, 1.02-13.05; P = 0.009), PCPC (OR, 3.98; 99% CI, 1.09-14.45; P = 0.006), and all mortality (OR, 9.2; 99% CI, 0.91-92.6; P = 0.01; default mode network with developmental delay (OR, 4.14; 99% CI, 1.19-14.43; P = 0.003); PCPC (OR, 4.1; 99% CI, 1.2-14.2; P = 0.004), NICU mortality (OR, 20.41; 99% CI, 0.89-468; P = 0.01), and motor tone/weakness (OR, 3.35; 99% CI, 1.01-11.12; P = 0.009); and seizure onset zone with concern for seizures (OR, 4.02; 99% CI, 1.0-16.15; P = 0.01). Of the other acute phase tests, only anatomical MRI was showed association with and outcome, concern for seizure (OR, 2.40; 99% CI, 0.94-6.13; P = 0.01).

**Conclusions:** This study provides level 3 evidence (OCEBM Levels of Evidence Working Group) demonstrating that in neonatal acute brain injury, the degree of abnormality of resting state networks is associated with mortality, ongoing concern for seizure and 2 year outcomes. These findings suggest RS is feasible and safe to implement in a busy tertiary neonatal ICU and the findings are of at least equivalent value to other standard of care diagnostics.

**Highlights:**

- Cognition is incrementally associated with the DMN, Lang/FP, and the BG at two years in neonatal ABI.
- Motor outcomes are incrementally associated with the BG and DMN at two years in neonatal ABI.
- Seizure outcomes are incrementally associated with rs-SOZ at two years in neonatal ABI.
- Mortality after discharge is incrementally associated with the DMN and BG.
- Compared to EEG, MRS, and task-fMRI, only anatomical MRI had 2-year association with outcomes, on-going concern for seizure at 2 years in neonatal ABI.
- RS is feasible and safe to implement in a busy tertiary neonatal ICU and the findings are of at least equivalent value to other standard of care diagnostics.

## 1. Introduction

Internationally, 2-6 per 1000 live births have acute brain injury (ABI), of which 1.5 per 1000 is from neonatal hypoxic ischemic encephalopathy (HIE) ^1,2^. In the post-cooling era HIE, there is 20% moderate to severe morbidity and 25% mortality, with 91% of deaths due to withdrawal of life sustaining therapy (WSLT) in one study ^3^. WLST is informed by DOC exam linked to brain imaging and electrophysiology ^4^. However, since there are no consensus definitions of neonatal consciousness or DOC ^5^, the link between exam and imaging-electrophysiology is inherently less sensitive and specific in neonates. If neonatal consciousness cannot be determined or at best encompasses less specific DOC categorizations than children and adults, then reliance on imaging-electrophysiology in WLST determinations will remain higher in neonates. Further adding to this challenge, the current standard implementation of neonatal imaging-electrophysiology does not commonly yet include the gold standard of biomarker of consciousness, integrated brain network function ^6-13^. Integrated brain network function measures are stimulation/task-based and resting state electroencephalogram (EEG) and functional magnetic resonance imaging. These tests are recommended to classify DOC and supplant the exam when the former indicate higher capacity for consciousness and predict outcomes in adults^14^.

In *healthy* neonates, resting state networks (RSNs) are detectable by the 3^rd^ trimester ^15^. The default mode (DMN), attention, and frontoparietal (FP) networks are detectable by term ^16,17^. Also by term, the reciprocal relationship between the DMN and dorsal attention network is present, thus, neonates ***<u>possess key features of the neural circuitry that enables integration of information across diverse sensory and high-order functional modules</u>***, ***<u>giving rise to conscious awareness</u>***. ^18^

Though there are many normal to mild neonatal ABI RS studies with outcomes ^15,19-31^, only two included the severe ABI. ^32,33^ The first, showed RS network (RSN) association with motor outcomes. ^34^ The second was the progenitor to the current study, and it evaluated specific RSN’s association with acute-period neurological/consciousness exam, mortality, other standard tests, and outcomes at mean age of 7 months post-gestation ^35^. This study found that at 7 months, the basal ganglia resting state network (BG) and seizure onset zone networks (RS-SOZ) were associated with motor outcomes. The Broad language/cognitive region networks were associated with developmental delay. Discharge with mortality was linked to default mode and language/cognitive networks. Exams were not linked to networks after multiple testing corrections. Lack of detection of all studied networks only occurred in those who did not survive.

Since an accurate and comprehensive test of integrated brain network function is needed for neonates during the acute brain injury period to inform on morbidity, we performed the same analysis extending the outcome period to about two years and added a standardized measure of development, the Pediatric Cerebral Performance Category Scale (PCPC) ^36,37^.

## 2. Methods

Study patients were 40 consecutive ABI neonates previously included in Boerwinkle et al. ^35^ who received clinical resting state fMRI (RS) with MRI during the acute period hospitalization of a suspected brain insult as part of clinical care between March 2018 and July 2019 at Phoenix Children’s Hospital (PCH).

### Data Collection

Data from the acute period were collected as described previously from the medical record ^35^, and follow up data were included through December 8, 2021. The PCH IRB approved this retrospective follow-up analysis (PCH IRB-20-331) and waived consent for this study.

Data collected from the medical record included: demographics, diagnosis, presentation, exams/test (neurological exam, consciousness, anatomical MRI (a-MRI), task-fMRI, RS, magnetic resonance spectroscopy (MRS), acute-period EEG, discharge condition, mortality, repeat insult, and follow-up outcomes (development, motor tone, strength, seizure) at 6.3 to 42 months of life. The extended follow-up outcomes also included a PCPC score.

Acute-period diagnosis, exam, acute test findings and extended outcome (not including PCPC score) were defined using ordinal scores based on reports from respective treating pediatric specialists as described in Boerwinkle et al. ^35^. Briefly, ordinal scores were assigned based on relevant chart notes and reports by two independent, blinded, study personnel. If disagreement occurred, a third research personnel was available. All acute-period scores remained the same from Boerwinkle et al. ^35^ and extended outcome scores were abstracted using the same methods used for outcome scores.

All acute-period measures (HIE severity, diagnoses, presentation, and neurological and consciousness exams, and initial EEG reads), except for discharge condition, were abstracted from documentation prior to the MRI with RS. Initial EEG reads were from after HIE and presentation severity determination. Since no additional tests or appointments were conducted for this study, some patients did not have all tests done (e.g. task-fMRI, EEG, MRS) because they were not clinically indicated. In these cases, patients with missing data points were excluded from the respective analysis. Patients had extended follow up past the outcomes in Boerwinkle et al. ^35^. Most had follow-up notes addressing outcome measures. In cases where specific information was not available/addressed in medical notes, this data was coded as missing.

### Measures

#### Acute-period diagnoses

As in the previous study ^35^, the Sarnat-criteria ^38^ were used to determine the presence of HIE from neonatologist final report documentation. The treating neonatologist’s documentation for the need of airway or ventilatory support, cardiovascular intervention and the duration of respective interventions, were used to categorize presentation severity/distress.

#### Neurological and consciousness exam

Neurological and consciousness exams were typically performed on the day of admission or next morning if admitted overnight. These exams were performed by the consulting neurocritical care provider. Repeat exams were documented if new events occurred (e.g. new weakness, worsened encephalopathy, concern for seizure, new abnormal EEG). The same ordinal scales were used to code this information as in the previous study. Additionally, repeat insult was documented to differentiate outcomes related to acute-period exams and outcomes related to an additional brain insult. Death was also recorded in patients who died prior to NICU discharge. In the previous study, there were no further deaths after discharge.

#### Acute period tests

Test safety, timing and anesthesia are described in detail in Boerwinkle et al., ^35^. Briefly, all participants were considered safe and stable to transport to the MR scanner for clinical MRI by the neonatologist and received continuous observation for the exams. Scan time was approximately 1 hr and there were no safety events related to scanning. Anesthesia was not in use during the time of scan, as per patient care plans. Movement was minimized by feeding and swaddling the infants. Multiple levels of ear protection were used (earplugs, earmuffs, MRI headphones). Awake patients were rescheduled as needed. As this is a retrospective study, no additional tests were performed outside of patients’ clinical care plans. In neonates with HIE, MRI with RS scanning was performed after a 72-h period of therapeutic hypothermia and medications for symptomatic relief of cold temperature. In neonates with non-HIE brain MRI indications, MRI with RS occurred at a more variable time according to clinical indication see ^35 for details^. EEG was recorded using a standard array of 19 leads and continuous video (cv-EEG). cv-EEG was interpreted by a pediatric epileptologist. Neonates suspected to have HIE started receiving cv-EEG on the first day of admission and it was removed the day of the MRI exam.

#### Outcome diagnoses and exams

Discharge condition was determined by the discharging neonatologist as described in Boerwinkle et al., ^35^. The extended outcome follow-up for development, motor-tone, and seizure were also determined using the same information and categorization procedures as in Boerwinkle et al., 2022, only at a later date and from clinical notes by patient providers within the patient medical record. The PCPC^36,37^; was the only additional measure used for the extended follow-up period. Death after discharge during extended follow-up was also documented. In the previous study, there were no deaths after NICU discharge in the more immediate follow up period.

#### PCPC score

The only measure added to this analysis compared to Boerwinkle et al., ^35^ was the PCPC. The scale includes typical cognitive ability for age (PCPC score 1), mild cognitive disability (2), moderate cognitive disability (3), severe cognitive disability (4), vegetative state or coma (5), and brain death (6). PCPC rating has been demonstrated to have good interrater reliability ^36^ If patients had a PCPC score recorded in a note at time of extended follow-up, this score was used, however for the majority of patients it was not available. For these patients, a PCPC score was assigned post-hoc as follows: two study personnel independently reviewed the patient medical record for provider documentation and classified a PCPC score based on available information. In cases of disagreement, a third person reviewed the case.

#### MRI acquisition

Details of MRI scan acquisition parameters are detailed in Boerwinkle et al 2022. MRI sequences were acquired on a 3 Tesla MR scanner (Ingenia, Phillips Healthcare, Best, The Netherlands). T1, T2, T2* (EPI), and diffusion-weighted (DWI) sequences with corresponding apparent diffusion maps were acquired. Additionally, MRS was acquired using a single voxel point resolved spectroscopy sequence localized to the white matter of the centrum semiovale and grey matter of the basal ganglia. Interpretation of a-MRI, DWI, and MRS was performed by the pediatric neuroradiologist.

Patients who remained still also received up to two 5-minute passive task fMRIs. The tasks were passive movement of the index finger by the MRI technician and a kaleidoscope visual pattern. Both task-fMRI sequences were block design. Images were processed using DynaSuite Neuro Software (Invivo Corporation, Gainesville, FL) using a standard pipeline previously described ^35^. Task-fMRI was interpreted by the pediatric neuroradiologist.

#### RS

Details of the RS MRI acquisition parameters and preprocessing are in Boerwinkle et al., ^35^. In brief, 600 volumes were acquired per patient in two 10-minute scans with a repetition time (TR) of 2000 ms. After preprocessing, the FMRIB Software Library tool (FSL) MELODIC ^39^ was used on each patient individually, entering each fMRI run separately ^38,40^. The MELODIC results were manually sorted first into noise vs neuronal signal. Neuronal signal ICs were then further sorted into known typical resting state networks (RSNs) such as motor, vision, default mode, et cetera, and atypical neuronal signal sources. Details for this procedure and cohort are described in Boerwinkle et al., ^35^ The specific RSNs evaluated for this and our previous study were: (1) the basal ganglia (BG); (2) the language/frontoparietal network (Lang/FP); (3) the default mode network (DMN); and (4) atypical (non-RSN) neuronal networks. Non-RSNs were further classified as to whether they were consistent with RS-SOZ networks seen in children with drug resistant epilepsy (DRE) ^40,41^. It was also noted whether patients had one or more typical RSNs not detected.

### Statistical analyses

The distribution of demographics, clinical factors, and follow up outcomes were summarized using descriptive statistics, and compared between RSN categories using the Fisher exact (FE). The magnitude of association between RSN category scores and factors/outcomes was quantified using odds ratio (OR) estimates and corresponding confidence intervals (CI) from ordinal logistic regression analyses (OLR), or multinomial logistic regression models (MLR) when the proportional odds assumption was not valid. Similar analyses examined association of clinical factors and outcomes with findings from EEG, a-MRI, task-fMRI, and MRS modalities. For OLR/MLR models, the modality predictor ordinal levels were fit as a continuous score, coded using 0,1, and 2 for normal, atypical, and not detected RSNs, respectively. The ordinal scores of 0,1,2, and 3 representing normal, mildly abnormal, moderately abnormal, and severely abnormal, respectively, were used for EEG, a-MRI, MRS, and task-fMRI tests. All statistical tests were 2-sided, with statistical significance and CI adjusted using a Bonferroni correction to account for multiple test comparisons for each modality (adjusted threshold of P<.01 for 5 comparisons per modality).

## 3. Results

Our previous study (Boerwinkle et al. ^35^) contained 40 patients, five of whom who survived to discharge but were since lost to follow up (LTFU) with demographics, exam, and non-RSN test classifications detailed in **Table 1**.Of the 35 who survived discharge and remained in follow-up, 68% were male, with mean (SD) gestational age 37.8 (2.6) weeks, and follow-up median age (interquartile range) 30.5 (23.6, 36.7) months of life. Mortality during the initial NICU hospitalization was 3 of 40, and outpatient mortality, occurring since then was 4 of the remaining 35 in follow-up.

**Table 1:** Demographics and Baseline Clinical Factors in Current Study Patients.

| Factor | Followed<br>(N=35) | Lost TFU<br>(N=5) | Total<br>(N=40) | P-value |
| --- | --- | --- | --- | --- |
| **Gestational Age** (weeks) |  |  |  |  |
| Mean (SD) | 37.6 (2.6) | 39.2 (1.4) | 37.8 (2.5) | 0.28 |
| Median (Q1, Q3) | 38.3 (36.0, 39.7) | 39.0 (37.9, 40.0) | 38.4 (36.7, 39.9) |  |
| **Sex**, N (%) |  |  |  |  |
| 0: Female | 14 (40) | 2 (40) | 16 (40) | 1.00 |
| 1: Male | 21 (60) | 3 (60) | 24 (60) |  |
| **HIE** |  |  |  |  |
| Yes, (N=27) |  |  |  |  |
| 1: Mild | 12 (50) | 2 (67) | 14 (52) | 1.00 |
| 2: Moderate | 6 (25) | 1 (33) | 7 (26) |  |
| 3: Severe | 6 (25) | 0 (0) | 6 (22) |  |
| 4: Unknown severity | 0 (0) | 0 (0) | 0 (0) |  |
| No, (N=13) |  |  |  |  |
| 1: Neonatal epilepsy | 3 (27) | 0 (0) | 3 (23) | 0.04 |
| 2: Genetic diagnosis or congenital malf. | 4 (36) | 0 (0) | 4 (31) |  |
| 3: Cardiac diagnosis | 0 (0) | 0 (0) | 0 (0) |  |
| 4: Infection | 0 (0) | 1 (50) | 1 (8) |  |
| 5: TBI | 2 (18) | 0 (0) | 2 (15) |  |
| 6: Hypoglycemia or similar metabolic derangement | 0 (0) | 1 (50) | 1 (8) |  |
| 7: Arterial or venous ischemic infarction | 2 (18) | 0 (0) | 2 (15) |  |
| **Presentation Severity**,<br>N (%) |  |  |  |  |
| 0: Normal | 1 (3) | 0 (0) | 1 (3) | 0.42 |
| 1: Mild distress | 16 (46) | 3 (60) | 19 (48) |  |
| 2: Moderate distress | 7 (20) | 2 (40) | 9 (23) |  |
| 3: Severe distress requiring pressors and prolonged ventilatory support | 11 (31) | 0 (0) | 11 (28) |  |
| **Neuro Exam Focalities**, N (%) |  |  |  |  |
| 0: Normal | 7 (20) | 3 (60) | 10 (25) | 0.45 |
| 1: Mild abnormal | 11 (31) | 1 (20) | 12 (30) |  |
| 2: Moderate abnormal | 12 (34) | 1 (20) | 13 (33) |  |
| 3: Severe abnormal | 5 (14) | 0 (0) | 5 (13) |  |
| **Acute State of Consciousness**<br>**(Day 0-5), N (%)**<br>0: Normal<br>1: Irritable or sleeping more but arouses easily and moves to touch normally<br>2: Difficult to wake up only to painful stimulation, and less spontaneous movements than typical<br>3: "Coma" = does not open eyes and does not move in response to environmental stimuli and painful stimulation or only abnormal movements<br>4: Episodically goes from normal to unresponsive = suggestive of possible seizure by bedside criteria | 7 (20)<br>13 (37)<br>11 (31)<br>3 (9)<br>1 (3) | 2 (40)<br>3 (60)<br>0 (0)<br>0 (0)<br>0 (0) | 9 (23)<br>16 (40)<br>11 (28)<br>3 (8)<br>1 (3) | 0.53 |
| **Anatomical MRI, N (%)**<br>0: Normal<br>1: Mild abnormal<br>2: Moderate abnormal<br>3: Severe abnormal | 12 (34)<br>9 (26)<br>5 (14)<br>9 (26) | 3 (60)<br>2 (40)<br>0 (0)<br>0 (0) | 15 (38)<br>11 (28)<br>5 (13)<br>9 (23) | 0.53 |
| **Task MRI, N (%)**<br>0: Normal<br>1: Mild abnormal<br>2: Moderate abnormal<br>3: Severe abnormal<br>4: Not done | 25 (71)<br>0 (0)<br>3 (9)<br>3 (9)<br>4 (11) | 5 (100)<br>0 (0)<br>0 (0)<br>0 (0)<br>0 (0) | 30 (75)<br>0 (0)<br>3 (8)<br>3 (8)<br>4 (10) | 1.00 |
| **MR Spec, N (%)**<br>0: Normal<br>1: Mild abnormal<br>2: Moderate abnormal<br>3: Severe abnormal<br>4: Not done | 9 (26)<br>4 (11)<br>1 (3)<br>3 (9)<br>18 (51) | 3 (60)<br>1 (20)<br>0 (0)<br>0 (0)<br>1 (20) | 12 (30)<br>5 (13)<br>1 (3)<br>3 (8)<br>19 (48) | 0.45 |
| **EEG, N (%)**<br>0: Normal<br>1: Mild background abn only<br>2: Seizure<br>3: Flat<br>4: Not done | 2 (6)<br>19 (54)<br>12 (34)<br>1 (3)<br>1 (3) | 1 (20)<br>3 (60)<br>1 (20)<br>0 (0)<br>0 (0) | 3 (8)<br>22 (55)<br>13 (33)<br>1 (3)<br>1 (3) | 0.57 |

RSN classifications by normal (N), detected with atypical features (A), and not detectable (ND) are reported in **Table 2**. Compared to those who remained in follow-up, those LTFU with non-HIE etiologies, had higher neonatal epilepsy etiology than the other diagnostic categories (P=0.04). However, this was only two patients, and there was no significant difference in demographics, presentation, exam, test results, nor outcomes. Age and outcomes at most recent follow-up visit are provided in **Table 3**.

**Table 2:** RSN Classification Results – Normal, Atypical, and Not Detected.

| Diagnostic | Followed<br>(N=35) | Lost TFU<br>(N=5) | Total<br>(N=40) | P-value |
| --- | --- | --- | --- | --- |
| **Age at RS (weeks)** |  |  |  |  |
| All (N=40) |  |  |  |  |
| Mean (SD) | 2.18 (2.60) | 1.29 (0.79) | 2.07 (2.46) |  |
| Median (Q1, Q3) | 1.14 (0.71, 2.29) | 1.29 (0.57, 2.00) | 1.14 (0.64, 2.14) | 0.71 |
| HIE (N=27) |  |  |  |  |
| Mean (SD) | 1.61 (2.07) | 1.29 (0.71) | 1.57 (1.96) |  |
| Median (Q1, Q3) | 0.93 (0.64, 1.36) | 1.2 (0.57, 2.00) | 1.14 (0.57, 1.43) | 0.73 |
| Non-HIE (N=13) |  |  |  |  |
| Mean (SD) | 3.44 (3.27) | 1.29 (1.21) | 3.11 (3.11) |  |
| Median (Q1, Q3) | 2.14 (0.86, 4.43) | 1.29 (0.43, 2.14) | 2.14 (0.86, 4.00) | 0.46 |
| **BG, N (%)** |  |  |  |  |
| 0: Normal | 8 (23) | 2 (40) | 10 (25) |  |
| 1: Detected with atypical features | 21 (60) | 3 (60) | 24 (60) | 0.65 |
| 2: Not detected | 6 (17) | 0 (0) | 6 (15) |  |
| **BG** |  |  |  |  |
| Mean (SD) | 0.94 (0.64) | 0.60 (0.55) | 0.90 (0.63) |  |
| Median (Q1, Q3) | 1 (1, 1) | 1 (0, 1) | 1 (0.5, 1) | 0.38 |
| **Language and/or F-P, N (%)** |  |  |  |  |
| 0: Normal | 18 (51) | 5 (100) | 23 (58) |  |
| 1: Detected with atypical features | 13 (37) | 0 (0) | 13 (33) | 0.19 |
| 2: Not detected | 4 (11) | 0 (0) | 4 (10) |  |
| **Language and/or F-P** |  |  |  |  |
| Mean (SD) | 0.60 (0.69) | 0 (0) | 0.53 (0.68) |  |
| Median (Q1, Q3) | 0 (0, 1) | 0 (0, 0) | 0 (0, 1) | 0.08 |
| **Default Mode, N (%)** |  |  |  |  |
| 0: Normal | 22 (63) | 5 (100) | 27 (68) |  |
| 1: Detected with atypical features | 8 (23) | 0 (0) | 8 (20) | 0.35 |
| 2: Not detected | 5 (14) | 0 (0) | 5 (13) |  |
| **Default Mode** |  |  |  |  |
| Mean (SD) | 0.51 (0.74) | 0 (0, 0) | 0.45 (0.71) |  |
| Median (Q1, Q3) | 0 (0, 1) | 0 (0, 0) | 0 (0, 1) | 0.20 |
| **Seizure Onset Zone or Abnormal Findings Concerning for Seizure, N (%)** |  |  |  |  |
| 0: Normal | 19 (54) | 5 (100) | 24 (60) |  |
| 1: Yes some concern for seizure | 7 (20) | 0 (0) | 7 (18) | 0.25 |
| 2: Highly concerning for seizure | 9 (26) | 0 (0) | 9 (23) |  |
| **Seizure Onset Zone or Abnormal Findings Concerning for Seizure** |  |  |  |  |
| Mean (SD) | 0.71 (0.86) | 0 (0) | 0.63 (0.84) |  |
| Median (Q1, Q3) | 0 (0, 2) | 0 (0, 0) | 0 (0, 1) | 0.1 |

P-values are from the Wilcoxon Sum-Rank test for the continuous predictors and from Fisher's Exact Test for categorical predictors comparing differences between follow up and lost to follow up.

**Table 3:** Outcomes at Most Recent Visit

| **Outcome** | **Outcomes**<br>**(N=35)** |
| --- | --- |
| **Age at Follow-up** (months)<br>Mean (SD)<br>Median (Q1, Q3)<br>(Deceased total N=7) | 28.2 (10.7)<br>30.5 (23.6,<br>36.7) |
| **Developmental delay**, N (%)<br>0: Normal<br>1: Mild delay<br>2: Moderate delay or focal finding on exam<br>3: Severe findings<br>4: Deceased | 11 (31)<br>3 (9)<br>10 (29)<br>4 (11)<br>7 (20) |
| **Motor-Tone**, N (%)<br>0: Normal<br>1: Mildly increased tone or weakness<br>2: Moderately increased tone or weakness<br>3: Severely increased tone or weakness<br>4: Deceased | 16 (46)<br>7 (20)<br>2 (6)<br>3 (9)<br>7 (20) |
| **Concern for seizure or on anti-seizure med**, N (%)<br>(Dead after discharge have outcomes other than 4)<br>0: No<br>1: Yes<br>4: Deceased prior to discharge | 17 (49)<br>14 (40)<br>4 (11) |
| **Additional Severe Brain Insult**, N (%)<br>(Dead after discharge have outcomes other than 4)<br>0: No known insult<br>1: Yes<br>4: Deceased prior to discharge | 31 (89)<br>1 (3)<br>3 (9) |
| **PCPC**, N (%)<br>1: Age appropriate<br>2: Mild disability<br>3: Moderate disability<br>4: Severe disability<br>6: Brain death/death | 11 (31)<br>4 (11)<br>9 (26)<br>4 (11)<br>7 (20) |
| **Deceased**, N (%)<br>0: No<br>1: Yes | 28 (80)<br>7 (20) |

There was consistency between the outcome measure’s percentages of the categorizations, normal v. not normal (but alive), and highest severity, and when compared to prior, were similar:

- developmental condition 31% v. 49% (prior 49% v. 43%); highest severity 11% (prior 13%),
- PCPC 31% v. 48% (not evaluated in prior); highest severity 9% (no prior measure),
- motor-tone 46% v. 35% (prior 46% v. 46%); highest severity 11% (prior 13%), and
- concern for seizure 49% v. 40% (prior 64% v. 36%).

Current study RSN and other tests associations with outcomes (developmental delay, motor tone, concern for seizure, and PCPC) are detailed in **eTable 1-8**, with those surviving multiple testing corrections, per network and diagnostic test, compared to the prior study in **Tables 4 and 5**. As tabulated, the RSNs had association with developmental delay, PCPC, motor tone/weakness, and mortality, whereas rs-SOZ was associated with ongoing concern for seizures at 2 years. Comparatively, of the other acute phase tests (a-MRI, passive task-fMRI, MRS, and EEG), only a-MRI was incrementally associated with an outcomes, which was concern for seizure (OR, 2.40; 99% CI, 0.94-6.13; P = 0.01), consistent with the prior study (**eTable 5-8)**. Of note, a significant association was identified between the EEG with concern for seizure within the 7-month period of the previous study but did not remain significant in the current. As prior, mortality had no significant association with a-MRI, task-fMRI, MRS, and EEG.

**Table 4.** Summary RSN association with Outcomes, showing only those associations with RSN degradation incrementally association with worsening outcomes ***<u>across the entire severity spectrum</u>***, and rs-SOZ associated with ongoing concern for seizure designated as YES or NO.

|  | Current | Prior |
| --- | --- | --- |
| **BG** | • PCPC (OR, 9.54; 99% CI, 1.89-48.1; $P = 0.0003$)<br>• NICU mortality (OR, 57.5; 99% CI, 1.35->999; $P = 0.006$)<br>• All mortality (OR, 65.7; 99% CI 1.47->999; $P = 0.005$)<br>• motor tone/weakness (OR, 17.8; 99% CI, 2.2-143; $P = 0.0004$) | • developmental delay (odds ratio [OR], 14.5; 99.4% confidence interval [CI], 2.00–105; $P < .001$)<br>• motor tone/weakness (OR, 9.98; 99.4% CI, 1.72–57.9; $P < .001$) |
| **Lang/FP** | • developmental delay (OR, 3.64; 99% CI, 1.02-13.05; $P = 0.009$)<br>• PCPC (OR, 3.98; 99% CI, 1.09-14.45; $P = 0.006$)<br>• All mortality (OR, 9.2; 99% CI, 0.91-92.6; $P = 0.01$) | • outpatient developmental delay (OR, 4.77; 99.4% CI, 1.21–18.7; $P = .002$)<br>• discharge condition (OR, 5.13; 99.4% CI, 1.22–21.5; $P = .002$) |
| **DMN** | • developmental delay (OR, 4.14; 99% CI, 1.19-14.43; $P = 0.003$)<br>• PCPC (OR, 4.1; 99% CI, 1.2-14.2; $P = 0.004$)<br>• NICU mortality (OR, 20.41; 99% CI, 0.89-468; $P = 0.01$)<br>• motor tone/weakness (OR, 3.35; 99% CI, 1.01-11.12; $P = 0.009$) | • discharge condition (OR, 3.72; 99.4% CI, 1.01–13.78; $P = .006$) and<br>• neurological exam ($P = .002$ (FE); OR, 11.8; 99.4% CI, 0.73–191; $P = .01$ (OLR)) |
| **Rs-SOZ** | • concern for seizures (OR, 4.02; 99% CI, 1.0-16.15; $P = 0.01$) | • motor tone/weakness (OR, 3.31; 99.4% CI, 1.08–10.1; $P = .003$) |

**Table 5.**
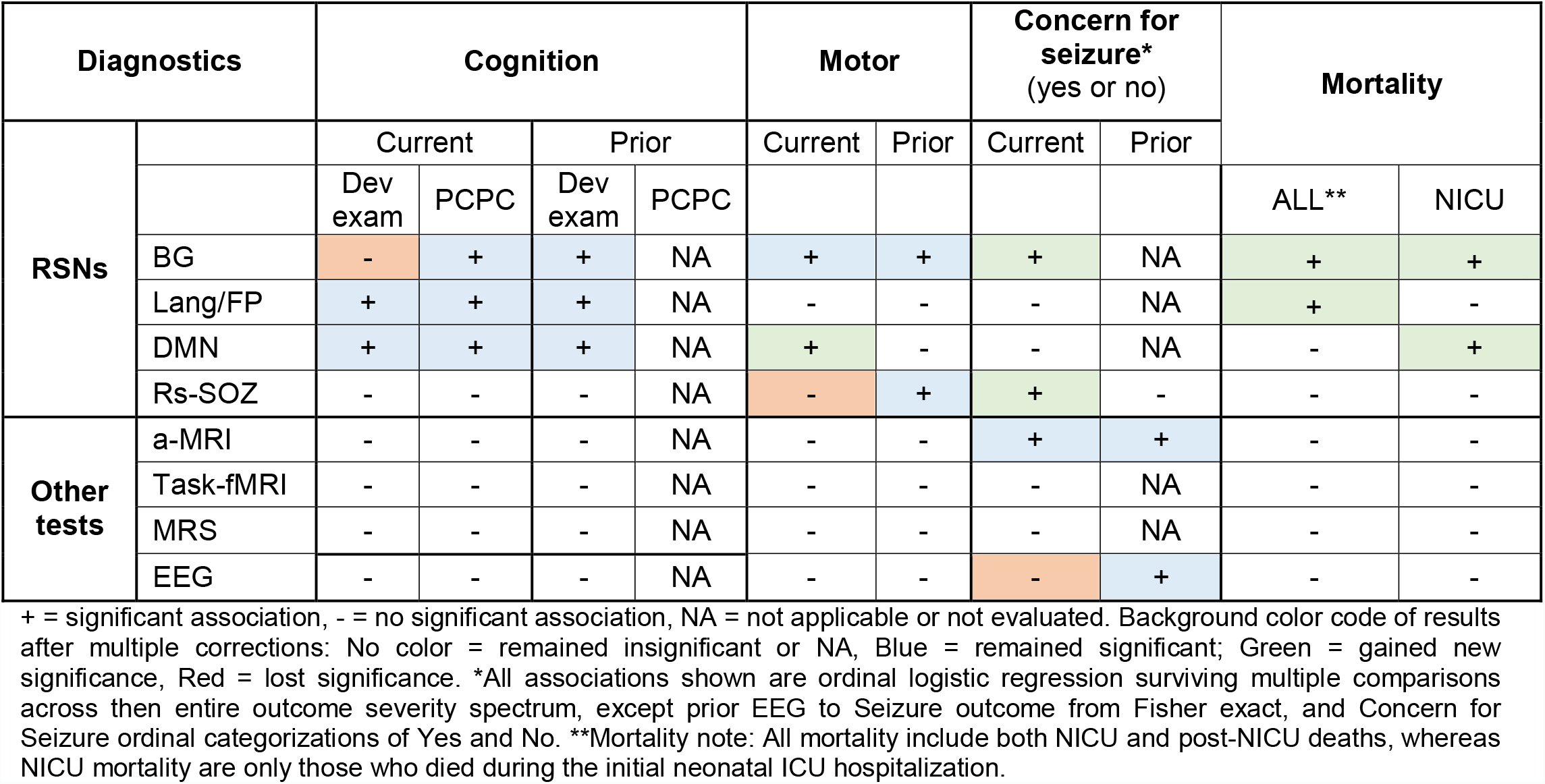
Comparison of Prior and Current Study Incremental Associations Across Entire Outcome Severity Spectrum

In overview, together RSNs and rs-SOZ gained associations with motor, concern for seizure, and all mortality, except rs-SOZ with motor. The RSNs all had association with PCPC, and remained stable in association with developmental delay, except the BG. Of the other tests, a-MRI-seizure remained stable in association, whereas EEG-seizure no longer met significant association.

Of note, between the prior and current study, NICU mortality was specifically included as an outcome in the correction for multiple comparisons, rather than as part of the several options of discharge condition in the prior study. Also, compared to prior, the RSN-concern for seizure was not evaluated, thus is it unknown if the association between it and BG is stable or gained, for simplicity is reported and newly determined/gained here.

According to the leading experts in coma and DOC, differentiating between WLST and other causes of death is critical to understand if a person could have been supported indefinitely or died, despite maximal medical therapy for the patient’s environment^4^. This distinction is one means by which discernment of RSN as a biomarker of integrated brain network function is related to a recoverable and sustainable neurological state with adequate quality of life. However, in neonatal ABI, not only is WLST distinction often not clearly documented, so is the lack of consensus definition of the adequate quality of life in the early life period. Ultimately, since our goal is to decide if it was the poor neurological function that possibly led to the cascade of events to death, rather than define WLST v non-WLST in a neonate, we determined the congruency between RS and events likely leading to death in the 7 mortalities. Six were congruent, wherein both the degree of abnormal RS networks and neurological diagnosis/events were most likely causative. The remaining case cause of death was due to organ failure deemed non-neurologically related (data supporting these outcomes are available by contacting the investigators).

## 4. Discussion

These RS findings, pointing to ABI impacting the neural network function and plasticity, are consistent with results from other modalities evaluating adults and children with ABI ^42^.

Thus, there are several indicators that this neonatal population reflects the severity of brain pathology expected from a tertiary referral center, who are the intended recipients of the RS. Seven of 40 (17.5%) patients died, and four were after initial discharge. 35 of 40 (87.5%) continued to care at the tertiary center over 2 years later. 49% had abnormal cognitive development, 35% abnormal motor outcomes, and 40% with concern for seizure, and around 10% of each outcome were in the most severe category, leaving only nearly equal numbers in the typical development range.

### 4.1 Cognition

Compared to the other diagnostics, the association between developmental delay and the RSNs, including the BG, Lang/FP, and DMN remained significant between the current and prior studies, except for the developmental exam and BG, though the PCPC was associated with the BG connectivity. This association pattern is integrally related to the PCPCs reflection of cognitive impairment.

It is worth recalling that the PCPC was developed as a means of quantifying cognitive impairment in children after critical illness or injury ^36^. The scale includes typical cognitive ability for age (PCPC score 1), mild cognitive disability (2), moderate cognitive disability (3), severe cognitive disability (4), vegetative state or coma (5), and brain death (6). PCPC rating has been demonstrated to have good interrater reliability ^36^. Validity of the scale was initially demonstrated relative to multiple measures of morbidity in critically ill infants and children. Specifically for infants, PCPC scores have been correlated with length of stay, total hospital charges, discharge care needs, and predicted mortality rate ^36^ – all relevant to the current study. PCPC has also been shown to have correlation with Stanford-Binet Intelligence Quotients and Bayley Mental Developmental Index scores ^37^. PCPC score has been utilized as an outcome measure in other neonatal follow up studies ^43^, and is frequently utilized in large pediatric neurocritical care research ^44^.

Supportively, FP is one of the early networks detected, as Doria et al. found FP present by term ^15^, and may be predictive of individually unique cognitive capacities ^45^. This may help explain the cognitive and other cortically localized outcomes known to occur after HIE, though longer follow up is indicated ^46-48^.

### 4.2 Motor & Tone

Compared to the other diagnostics, the association between motor and tone development and the BG and DMN, remained or became significant, respectively, between the prior and current studies – with the exception of rs-SOZ which was no longer associated. The consistent lack and loss of relationship between Lang/FP and rs-SOZ, respectively with motor development, in the context of continued BG association is the first network-specific prognostic to a single developmental stream in humans.

In clinical practice, broad ranges of developmental outcomes are often presented to families of neonates with ABI, lumping developmental streams together. In addition to the social-emotional costs to the family, the medical and societal costs of applying equivalent resources with the current lack of specifically are untold. By applying specific biomarkers to separate developmental streams such as herein with the BG RSN and motor outcome, we may increase our neuro-prognosticative specificity and effectiveness of resultant resource utilization. Compared to other prior studies the BG network relationship to motor pathology and development in neonates is consistent (Linke et al ^30^).

### 4.3 Seizure and Epileptogenesis

Unexpectedly, association with concern for seizure was gained by rs-SOZ gained, whereas this relationship was lost with acute-phase EEG. This speaks toward rs-SOZ being more of a marker for epileptogenesis than acute-phase neonatal EEG. The relationship with anatomical MRI and concern for seizure remained significant, speaking toward this diagnostics relationship to both acute phase seizures and ongoing epileptogenesis, which may be due to the relationship between anatomic injury to the brain and role this plays in encouraging imbalance in excitation relative to suppression in network dynamics. Compared to other prior studies the rs-SOZ network relationship to epileptogensis and seizures in children and adults is consistent ^33,35,40,49^.

### 4.4 Coma and Mortality

Coma and other disorders of consciousness have high mortality, especially in the acute phase of brain injury. In the prior study, we showed that DMN associated with the acute neonatal neurologic exam, which included evaluation of consciousness, defined by responsiveness and eye opening to environmental stimuli. This network outcome association is supported by prior work ^13,50^ in other ABI populations. Comparatively, a-MRI was the only non-RSN test also associated with the neurological exam, similarly demonstrating its value in the post-cooling era ^51^.

The prior study showed lack of all studied networks only occurring in those who did not survive. In the current study, mortality remained associated with the BG, and gains connectivity association significance with the DMN. Whereas, before this DMN association did not survive multiple comparisons. Our cohort of neonates were term, a gestational age in which the reciprocal DMN-dorsal attention network is present, which enables integration of information across diverse sensory and high-order functional modules, giving rise to conscious awareness ^18^. Thus, explaining the connection between the DMN and NICU mortality. Incremental degradation of this connectivity increased risk of NICU death for both the BG and DMN, but also as an outpatient in relation to the BG. Such outpatient relationship between BG and mortality is likely due to the abnormal motor-tone effect on respiratory and airway safety and stability and known effect of cerebral palsy.

Overall, the RS network abnormalities were congruent in all mortalities.

### 4.5 Limitations and Further Study

A prospective approach with standardized acute exams, and definition of neonatal consciousness and disorders of consciousness are needed. Our preliminary data suggests that the burden of common multiple atypical findings such as lack of recognizable detection of key modulating and cognition networks in individual neonates may cluster in those with the worst outcomes and is of great interest for future study. Clinical RS requires interpretation, thus the blinded study design, however automated RSN categorization would reduce bias ^52-54^.

In neonates with ABI, network pathology characterization would improve with further validation. Data-driven approaches, such as ICA, may best be suited to also minimize bias. ICA has level 1 evidence for diagnostic testing (OCEBM Levels of Evidence Working Group) in children and adults for characterizing normal and pathological networks in DRE and improving outcomes ^54^. Method comparison research may be of value.

There were fewer multiple comparison in the current study compared to prior, because in the prior we evaluated acute exam findings, which may be why significance varied.

## Conclusions

This study provides level 3 evidence (OCEBM Levels of Evidence Working Group) that RSN network connectivity degradation is associated with incrementally worsening severity of longitudinal and 2-year outcomes in mild to severe neonatal ABI. Two-year cognition, measured by the PCPC, was incrementally associated with the DMN, Lang/FP, and the BG. The BG and rs-SOZ were incrementally associated with 2-year motor outcomes and ongoing concern for seizure, respectively. The DMN and BG connectivity were associated with 2-year outpatient mortality. Comparatively, of the acute phase anatomical MRI, EEG, MRS, and task-fMRI, only anatomical MRI had association with outcomes at the 2-year mark, which was on-going concern for seizure. These findings suggest RS is feasible and safe to implement in a busy tertiary neonatal ICU and the findings are of at least equivalent value to other standard of care diagnostics.

This research did not receive any specific grant from funding agencies in the public, commercial, or not-for-profit sectors.

## Data Availability

All data produced in the present work are contained in the manuscript

## Abbreviations

LTFU: lost to follow up
ABI: acute brain injury
DOC: disorders of consciousness
EEG: electroencephalogram
fMRI: functional magnetic resonance imaging
RSN: resting state network(s)
DMN: default mode network
FP: frontoparietal network
Lang/FP: language/frontoparietal network
HIE: hypoxic ischemic encephalopathy
a-MRI: anatomical MRI
RS: resting state functional magnetic resonance imaging
MRS: magnetic resonance spectroscopy
cv-EEG: continuous video EEG
ICA: independent component analysis
BOLD: blood oxygenation level dependent signal
DRE: drug resistant epilepsy
FE: Fisher exact
OR: odds ratio
CI: confidence intervals
OLR: ordinal logistic regression analyses
MLR: multinomial logistic regression models
CC: correlation coefficient
TBI: traumatic brain injury
RS-SOZ: seizure onset zone networks
PCPC: Pediatric Cerebral Performance Category Scale
BG: basal ganglia resting state network
DWI: diffusion-weighted imaging
IC: -------
TR: repetition time
DRE: drug resistant epilepsy
WLST: withdrawal of life sustaining therapies
non-RSN: atypical neuronal networks
NICU: neonatal intensive care unit

## Supplementary Materials

**eTable 1-4:** * P value significant at <.05 (not adjusted using Bonferroni correction). ** P value significant or marginally significant at <.01 (adjusted using Bonferroni correction). ^a^ P value from Fisher exact test; ^b^ P value from ordinal/multinomial logistic regression.

**eTable 1:**
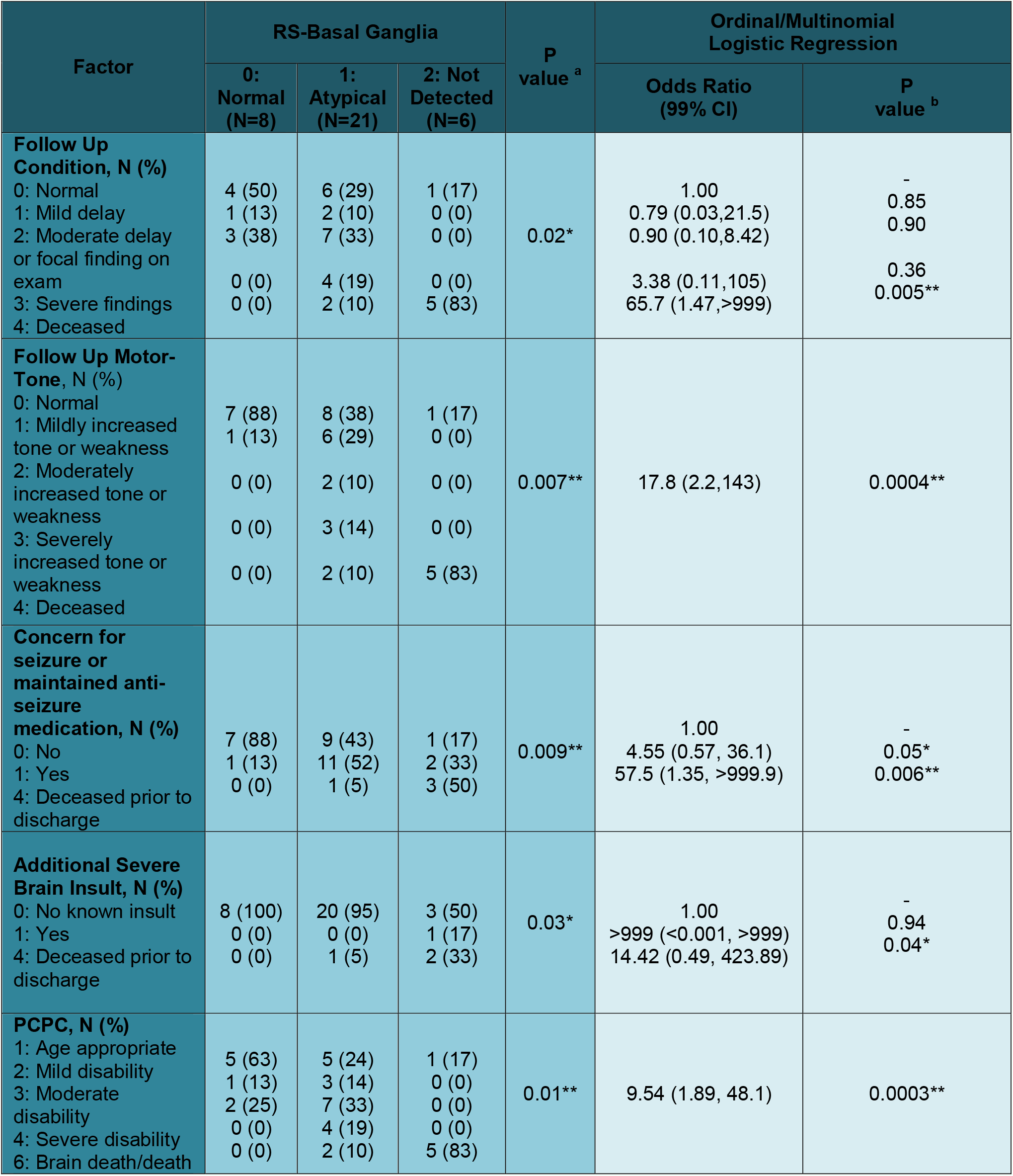
Association of Resting State Basal Ganglia (RS-BG) with Outcomes.

| Factor | RS-Basal Ganglia |  |  | P value <sup>a</sup> | Ordinal/Multinomial Logistic Regression |  |
| --- | --- | --- | --- | --- | --- | --- |
|  | 0: Normal (N=8) | 1: Atypical (N=21) | 2: Not Detected (N=6) |  | Odds Ratio (99% CI) | P value <sup>b</sup> |
| **Follow Up Condition, N (%)** |  |  |  |  |  |  |
| 0: Normal | 4 (50) | 6 (29) | 1 (17) | 0.02* | 1.00 | - |
| 1: Mild delay | 1 (13) | 2 (10) | 0 (0) |  | 0.79 (0.03,21.5) | 0.85 |
| 2: Moderate delay or focal finding on exam | 3 (38) | 7 (33) | 0 (0) |  | 0.90 (0.10,8.42) | 0.90 |
| 3: Severe findings | 0 (0) | 4 (19) | 0 (0) |  | 3.38 (0.11,105) | 0.36 |
| 4: Deceased | 0 (0) | 2 (10) | 5 (83) |  | 65.7 (1.47,>999) | 0.005** |
| **Follow Up Motor-Tone, N (%)** |  |  |  |  |  |  |
| 0: Normal | 7 (88) | 8 (38) | 1 (17) | 0.007** | 17.8 (2.2,143) | 0.0004** |
| 1: Mildly increased tone or weakness | 1 (13) | 6 (29) | 0 (0) |  |  |  |
| 2: Moderately increased tone or weakness | 0 (0) | 2 (10) | 0 (0) |  |  |  |
| 3: Severely increased tone or weakness | 0 (0) | 3 (14) | 0 (0) |  |  |  |
| 4: Deceased | 0 (0) | 2 (10) | 5 (83) |  |  |  |
| **Concern for seizure or maintained anti-seizure medication, N (%)** |  |  |  |  |  |  |
| 0: No | 7 (88) | 9 (43) | 1 (17) | 0.009** | 1.00 | - |
| 1: Yes | 1 (13) | 11 (52) | 2 (33) |  | 4.55 (0.57, 36.1) | 0.05* |
| 4: Deceased prior to discharge | 0 (0) | 1 (5) | 3 (50) |  | 57.5 (1.35, >999.9) | 0.006** |
| **Additional Severe Brain Insult, N (%)** |  |  |  |  |  |  |
| 0: No known insult | 8 (100) | 20 (95) | 3 (50) | 0.03* | 1.00 | - |
| 1: Yes | 0 (0) | 0 (0) | 1 (17) |  | >999 (<0.001, >999) | 0.94 |
| 4: Deceased prior to discharge | 0 (0) | 1 (5) | 2 (33) |  | 14.42 (0.49, 423.89) | 0.04* |
| **PCPC, N (%)** |  |  |  |  |  |  |
| 1: Age appropriate | 5 (63) | 5 (24) | 1 (17) | 0.01** | 9.54 (1.89, 48.1) | 0.0003** |
| 2: Mild disability | 1 (13) | 3 (14) | 0 (0) |  |  |  |
| 3: Moderate disability | 2 (25) | 7 (33) | 0 (0) |  |  |  |
| 4: Severe disability | 0 (0) | 4 (19) | 0 (0) |  |  |  |
| 6: Brain death/death | 0 (0) | 2 (10) | 5 (83) |  |  |  |

**eTable 2:**
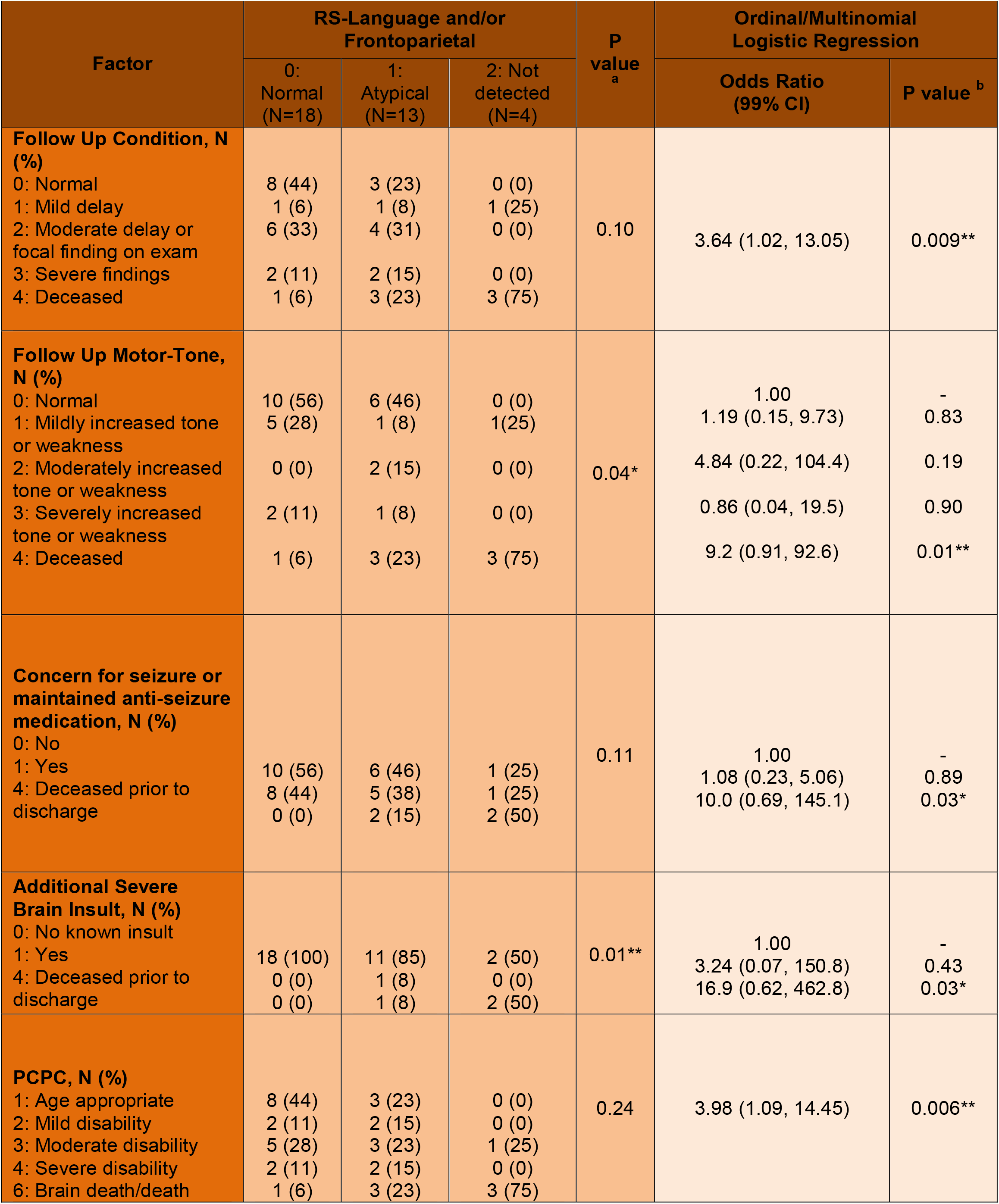
Association of Resting State (RS) Language and/or Frontoparietal (Lang-FP) with Outcomes.

**eTable 3:**
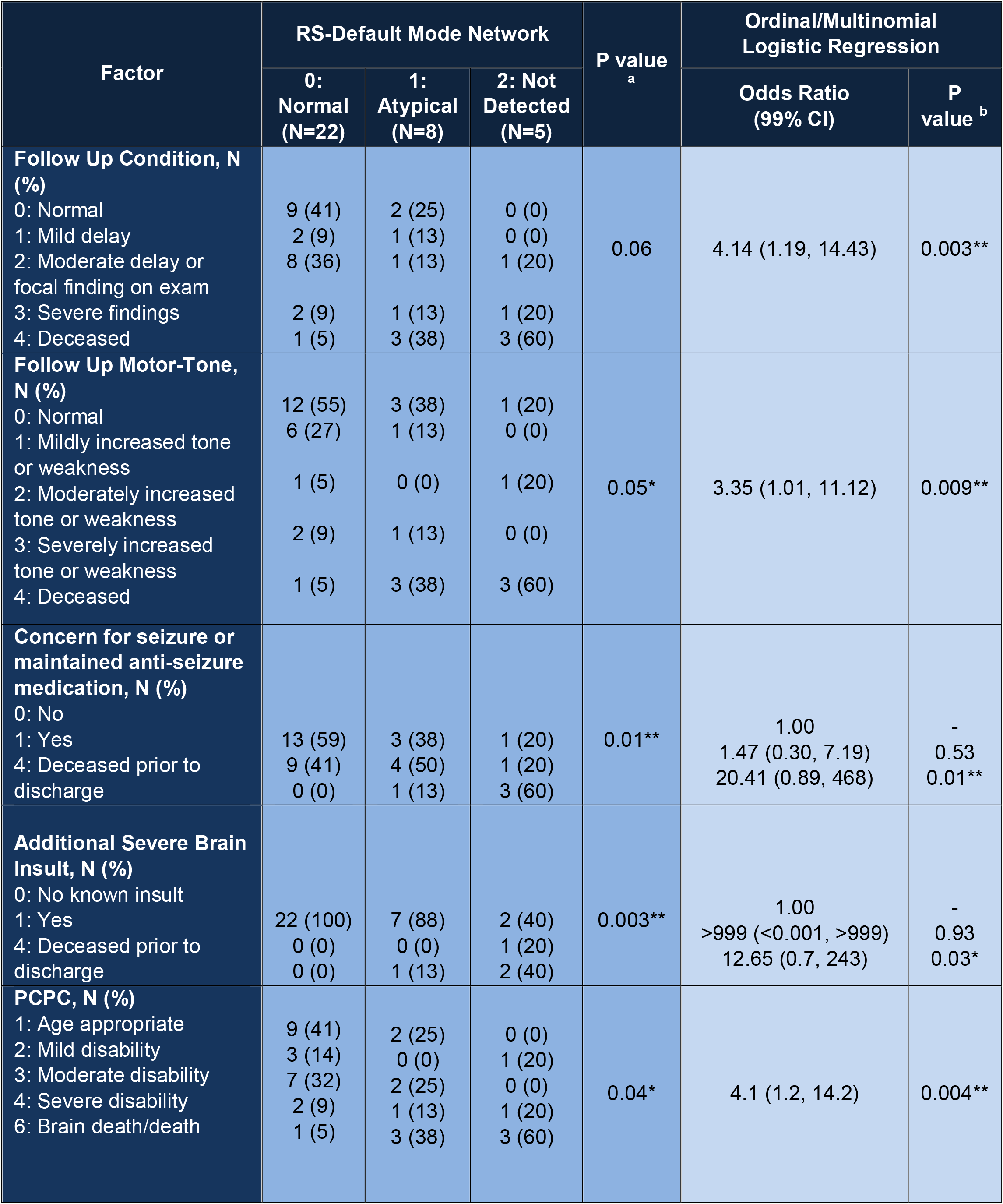
Association of Resting State (RS) Default Mode Network with Outcomes.

**eTable 4:**
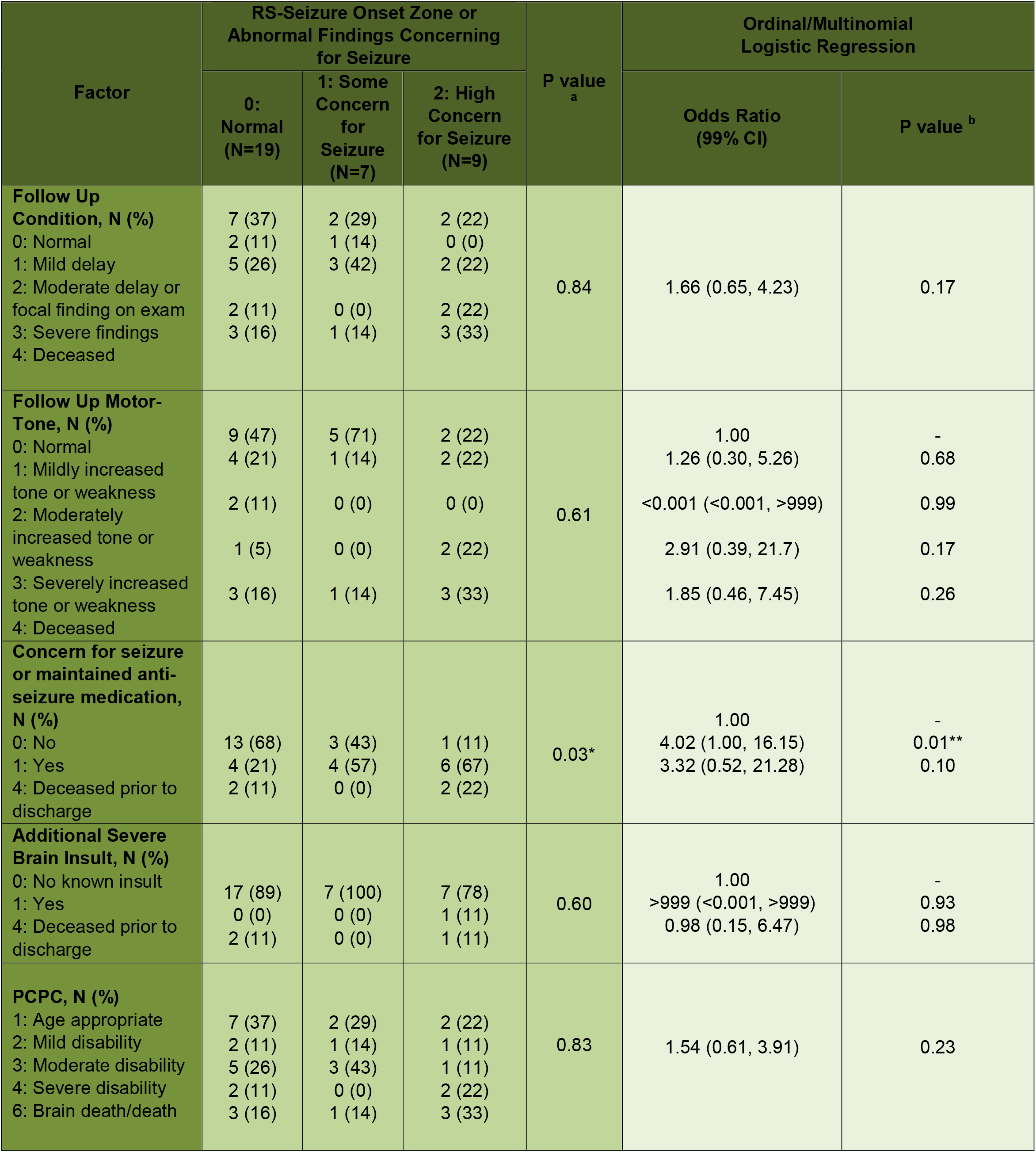
Association of Resting State (RS) Seizure Onset Zone or Abnormal Findings Concerning for Seizure with Outcomes.

**eTable 5-8:** * P value significant at <.05 (not adjusted using Bonferroni correction). ** P value significant or marginally significant at <.01 (adjusted using Bonferroni correction). ^a^ P value from Fisher exact test; ^b^ P value from ordinal/multinomial logistic regression.

**eTable 5:** Association of Baseline Factors with Anatomical MRI.

| Factor | Anatomical MRI |  |  |  | P value <sup>a</sup> | Ordinal/Multinomial Logistic Regression |  |
| --- | --- | --- | --- | --- | --- | --- | --- |
|  | 0: Normal (N=12) | 1: Mildly Abnormal (N=9) | 2: Moderately Abnormal (N=5) | 3: Severely Abnormal (N=9) |  | Odds Ratio (99% CI) | P value <sup>b</sup> |
| **Follow Up Condition, N (%)**<br>0: Normal<br>1: Mild delay<br>2: Moderate delay or focal finding on exam<br>3: Severe findings<br>4: Deceased | 5 (42)<br>1 (8)<br>3 (25)<br>1 (8)<br>2 (17) | 4 (44)<br>0 (0)<br>2 (22)<br>1 (11)<br>2 (22) | 1 (20)<br>1 (20)<br>1 (20)<br>2 (40)<br>0 (0) | 1 (11)<br>1 (11)<br>4 (44)<br>0 (0)<br>3 (33) | 0.57 | 1.38 (0.71, 2.69) | 0.21 |
| **Follow Up Motor-Tone, N (%)**<br>0: Normal<br>1: Mildly increased tone or weakness<br>2: Moderately increased tone or weakness<br>3: Severely increased tone or weakness<br>4: Deceased | 7 (58)<br>2 (17)<br>1 (8)<br>0 (0)<br>2 (17) | 4 (44)<br>2 (22)<br>0 (0)<br>1 (11)<br>2 (22) | 1 (20)<br>2 (40)<br>0 (0)<br>2 (40)<br>0 (0) | 4 (44)<br>1 (11)<br>1 (11)<br>0 (0)<br>3 (33) | 0.49 | 1.31 (0.67, 2.59) | 0.30 |
| **Concern for seizure or maintained anti-seizure medication, N (%)**<br>0: No<br>1: Yes<br>4: Deceased prior to discharge | 9 (75)<br>2 (17)<br>1 (8) | 5 (56)<br>3 (33)<br>1 (11) | 1 (20)<br>4 (80)<br>0 (0) | 2 (22)<br>5 (56)<br>2 (22) | 0.12 | 1.00<br>2.40 (0.94, 6.13)<br>2.22 (0.59, 8.27) | -<br>0.01**<br>0.12 |
| **Additional Severe Brain Insult, N (%)**<br>0: No known insult<br>1: Yes<br>4: Deceased prior to discharge | 11 (92)<br>1 (8)<br>0 (0) | 8 (89)<br>0 (0)<br>1 (11) | 5 (100)<br>0 (0)<br>0 (0) | 7 (78)<br>0 (0)<br>2 (22) | 0.49 | 1.00<br><0.001 (<0.001, >999)<br>2.33 (0.47, 11.6) | -<br>0.90<br>0.18 |
| **PCPC, N (%)**<br>1: Age appropriate<br>2: Mild disability<br>3: Moderate disability<br>4: Severe disability<br>6: Brain death/death | 6 (50)<br>1 (8)<br>2 (17)<br>1 (8)<br>2 (17) | 3 (33)<br>1 (11)<br>2 (22)<br>1 (11)<br>2 (22) | 1 (20)<br>1 (20)<br>1 (20)<br>2 (40)<br>0 (0) | 1 (11)<br>1 (11)<br>4 (44)<br>0 (0)<br>3 (33) | 0.59 | 1.49 (0.76, 2.93) | 0.13 |

**eTable 6:** Association of Baseline Factors with Task-fMRI.

| Factor | Task-fMRI |  |  | P value <sup>a</sup> | Ordinal/Multinomial Logistic Regression |  |
| --- | --- | --- | --- | --- | --- | --- |
|  | 0: Normal (N=25) | 2: Moderately Abnormal (N=3) | 3: Severely Abnormal (N=3) |  | Odds Ratio (99% CI) | P value <sup>b</sup> |
| **Follow Up Condition, N (%)**<br>0: Normal<br>1: Mild delay<br>2: Moderate delay or focal finding on exam<br>3: Severe findings<br>4: Deceased | 8 (32)<br>2 (8)<br>6 (24)<br>4 (16)<br>5 (20) | 0 (0)<br>1 (33)<br>2 (67)<br>0 (0)<br>0 (0) | 0 (0)<br>0 (0)<br>1 (33)<br>0 (0)<br>2 (67) | 0.29 | 1.47 (0.64, 3.40) | 0.24 |
| **Follow Up Motor-Tone, N (%)**<br>0: Normal<br>1: Mildly increased tone or weakness<br>2: Moderately increased tone or weakness<br>3: Severely increased tone or weakness<br>4: Deceased | 11 (44)<br>5 (20)<br>1 (4)<br>3 (12)<br>5 (20) | 2 (67)<br>1 (33)<br>0 (0)<br>0 (0)<br>0 (0) | 0 (0)<br>0 (0)<br>1 (33)<br>0 (0)<br>2 (67) | 0.25 | 1.41 (0.61, 3.24) | 0.29 |
| **Concern for seizure or maintained anti-seizure medication, N (%)**<br>0: No<br>1: Yes<br>4: Deceased prior to discharge | 14 (56)<br>8 (32)<br>3 (12) | 1 (20)<br>4 (80)<br>0 (0) | 0 (0)<br>2 (67)<br>1 (33) | 0.23 | 1.00<br>2.36 (0.60, 9.26)<br>2.23 (0.44, 11.45) | -<br>0.10<br>0.21 |
| **Additional Severe Brain Insult, N (%)**<br>0: No known insult<br>1: Yes<br>4: Deceased | 22 (88)<br>1 (4)<br>2 (8) | 3 (100)<br>0 (0)<br>0 (0) | 2 (67)<br>0 (0)<br>1 (33) | 0.60 | 1.00<br><0.001 (<0.001, >999)<br>1.52 (0.43, 5.36) | -<br>0.99<br>0.39 |
| **PCPC, N (%)**<br>1: Age appropriate<br>2: Mild disability<br>3: Moderate disability<br>4: Severe disability<br>6: Brain death/death | 8 (32)<br>2 (8)<br>6 (24)<br>4 (16)<br>5 (20) | 0 (0)<br>1 (33)<br>2 (67)<br>0 (0)<br>0 (0) | 0 (0)<br>0 (0)<br>1 (33)<br>0 (0)<br>2 (67) | 0.27 | 1.47 (0.64, 3.40) | 0.24 |

**eTable 7:** Association of Baseline Factors with Magnetic Resonance Spectroscopy (MRS).

| Factor | MRS |  |  |  | P value <sup>a</sup> | Ordinal/Multinomial Logistic Regression |  |
| --- | --- | --- | --- | --- | --- | --- | --- |
|  | 0: Normal (N=9) | 1: Mildly Abnormal (N=4) | 2: Moderately Abnormal (N=1) | 3: Severely Abnormal (N=3) |  | Odds Ratio (99% CI) | P value <sup>b</sup> |
| **Follow Up Condition, N (%)**<br>0: Normal<br>1: Mild delay<br>2: Moderate delay or focal finding on exam<br>3: Severe findings<br>4: Deceased | 5 (56)<br>0 (0)<br>2 (22)<br>1 (11)<br>1 (11) | 2 (50)<br>0 (0)<br>1 (25)<br>1 (25)<br>0 (0) | 0 (0)<br>1 (100)<br>0 (0)<br>0 (0)<br>0 (0) | 0 (0)<br>0 (0)<br>2 (67)<br>0 (0)<br>1 (33) | 0.27 | 1.00<br>5.23 (0.28, 98.1)<br>3.49 (0.44, 27.73)<br>1.55 (0.09, 25.4)<br>3.72 (0.35, 40.2) | -<br>0.15<br>0.12<br>0.69<br>0.15 |
| **Follow Up Motor-Tone, N (%)**<br>0: Normal<br>1: Mildly increased tone or weakness<br>2: Moderately increased tone or weakness<br>3: Severely increased tone or weakness<br>4: Deceased | 5 (56)<br>2 (22)<br>0 (0)<br>1 (11)<br>1 (11) | 3 (75)<br>0 (0)<br>0 (0)<br>1 (25)<br>0 (0) | 1 (100)<br>0 (0)<br>0 (0)<br>0 (0)<br>0 (0) | 1 (33)<br>0 (0)<br>1 (33)<br>0 (0)<br>1 (33) | 0.73 | 1.00<br><0.001 (<0.001, >999)<br>>999 (<0.001, >999)<br>0.70 (0.06, 7.77)<br>1.71 (0.31, 9.59) | -<br>0.99<br>0.99<br>0.70<br>0.42 |
| **Concern for seizure or maintained anti-seizure medication, N (%)**<br>0: No<br>1: Yes<br>4: Deceased prior to discharge | 7 (78)<br>1 (11)<br>1 (11) | 2 (50)<br>2 (50)<br>0 (0) | 1 (100)<br>0 (0)<br>0 (0) | 0 (0)<br>2 (67)<br>1 (33) | 0.13 | 1.00<br>2.97 (0.61, 14.49)<br>2.78 (0.40, 19.42) | -<br>0.08<br>0.17 |
| **Additional Severe Brain Insult, N (%)**<br>0: No known insult<br>1: Yes<br>4: Deceased prior to discharge | 8 (89)<br>1 (11)<br>0 (0) | 4 (100)<br>0 (0)<br>0 (0) | 1 (100)<br>0 (0)<br>0 (0) | 2 (67)<br>0 (0)<br>1 (33) | 0.47 | 1.00<br><0.001 (<0.001, >999)<br>896 (<0.001, >999) | -<br>0.93<br>0.87 |
| **PCPC, N (%)**<br>1: Age appropriate<br>2: Mild disability<br>3: Moderate disability<br>4: Severe disability<br>6: Brain death/death | 4 (44)<br>1 (11)<br>2 (22)<br>1 (11)<br>1 (11) | 2 (50)<br>0 (0)<br>1 (25)<br>1 (25)<br>0 (0) | 0 (0)<br>0 (0)<br>1 (100)<br>0 (0)<br>0 (0) | 0 (0)<br>1 (33)<br>1 (33)<br>0 (0)<br>1 (33) | 0.86 | 1.51 (0.55, 4.18) | 0.29 |

**eTable 8:** Association of Baseline Factors with electroencephalogram (EEG).

| Factor | EEG |  |  |  | P value <sup>a</sup> | Ordinal/Multinomial Logistic Regression |  |
| --- | --- | --- | --- | --- | --- | --- | --- |
|  | 0: Normal (N=2) | 1: Mild Background Abnormality Only (N=19) | 2: Seizure (N=12) | 3: Flat (N=1) |  | Odds Ratio (99% CI) | P value <sup>b</sup> |
| **Follow Up Condition, N (%)**<br>0: Normal<br>1: Mild delay<br>2: Moderate delay or focal finding on exam<br>3: Severe findings<br>4: Deceased | 0 (0)<br>1 (50)<br>0 (0)<br>1 (50)<br>0 (0) | 8 (42)<br>1 (5)<br>5 (26)<br>2 (11)<br>3 (16) | 3 (25)<br>1 (8)<br>4 (33)<br>1 (8)<br>3 (25) | 0 (0)<br>0 (0)<br>0 (0)<br>0 (0)<br>1 (100) | 0.43 | 1.71 (0.48, 6.07) | 0.28 |
| **Follow Up Motor-Tone, N (%)**<br>0: Normal<br>1: Mildly increased tone or weakness<br>2: Moderately increased tone or weakness<br>3: Severely increased tone or weakness<br>4: Deceased | 1 (50)<br>0 (0)<br>0 (0)<br>1 (50)<br>0 (0) | 10 (53)<br>4 (21)<br>1 (5)<br>1 (5)<br>3 (16) | 5 (42)<br>2 (17)<br>1 (8)<br>1 (8)<br>3 (25) | 0 (0)<br>0 (0)<br>0 (0)<br>0 (0)<br>1 (100) | 0.73 | 1.93 (0.51, 7.28) | 0.20 |
| **Concern for seizure or maintained anti-seizure medication, N (%)**<br>0: No<br>1: Yes<br>4: Deceased prior to discharge | 1 (50)<br>1 (50)<br>0 (0) | 12 (63)<br>5 (26)<br>2 (11) | 3 (25)<br>8 (67)<br>1 (8) | 0 (0)<br>0 (0)<br>1 (100) | 0.07 | 1.00<br>2.92 (0.52, 16.45)<br>5.60 (0.43, 73.10) | -<br>0.11<br>0.08 |
| **Additional Severe Brain Insult, N (%)**<br>0: No known insult<br>1: Yes<br>4: Deceased prior to discharge | 2 (100)<br>0 (0)<br>0 (0) | 17 (90)<br>1 (5)<br>1 (5) | 11 (92)<br>0 (0)<br>1 (8) | 0 (0)<br>0 (0)<br>1 (100) | 0.23 | 1.00<br>0.42 (0.004, 40)<br>6.41 (0.34, 122) | -<br>0.63<br>0.10 |
| **PCPC, N (%)**<br>1: Age appropriate<br>2: Mild disability<br>3: Moderate disability<br>4: Severe disability<br>6: Brain death/death | 1 (50)<br>0 (0)<br>0 (0)<br>1 (50)<br>0 (0) | 7 (37)<br>3 (16)<br>4 (21)<br>2 (11)<br>3 (16) | 3 (25)<br>1 (8)<br>4 (33)<br>1 (8)<br>3 (25) | 0 (0)<br>0 (0)<br>0 (0)<br>0 (0)<br>1 (100) | 0.82 | 2.09 (0.57, 7.61) | 0.14 |

## Notes

### Competing Interest Statement

The authors have declared no competing interest.

### Funding Statement

This study did not receive any funding

### Author Declarations

The PCH IRB approved this retrospective follow-up analysis (PCH IRB-20-331) and waived consent for this study.

